# A clustering approach based on neuropsychological testing to detect early signs of probable MCI among community-dwelling older adults

**DOI:** 10.64898/2026.07.04.26357270

**Authors:** Rafael Mauti, Chiraz Chouk, Aymeric Guillot, Arnaud Peronin, Patrick Fargier, Pascal Chabaud

**Author notes:** co-last author (Patrick Fargier, Pascal Chabaud). **Correspondence:** Patrick Fargier.

## Abstract

**Introduction:** Mild Cognitive Impairment (MCI) is defined as a decline in one or more cognitive domains worse than expected for age and encompasses several subtypes depending on the cognitive(s) function(s) impaired. Older adults presenting amnestic MCI combined with one or several other impaired domains are the highest at risk of dementia. Yet, no consensual screening method for MCI in community-dwelling older adults has been established. The aim of this study was to identify cognitive aging profiles using multivariate analysis based on three neuropsychological assessments and to characterize profiles consistent with different probable MCI subtypes.

**Methods:** A total of 161 community-dwelling older adults from the 13EVAL cluster-randomized controlled trial performed the Montreal Cognitive Assessment including the Memory Index Score, the Trail Making Test and the Victoria Stroop Test. Hierarchical clustering on principal components was conducted based on four cognitive domains (global cognition, memory, inhibition and cognitive flexibility) and age. Inter-groups comparisons were performed using one-way ANOVA. Individual performances were compared to normative standard for each assessment to characterize each group.

**Results:** Three clusters were identified that differed significantly in age (*F*_2,158_=78.56, *p*<.001, *η^2^_p_* =.50), global cognition (*F*_2,158_=96.07, *p*<.001, *η^2^_p_* =.55), memory (*F*_2,158_=81.48, *p*<.001, *η^2^_p_*=.51), cognitive flexibility (*F*_2,158_=52.25, *p*<.001, *η^2^_p_* =.40) and inhibition (*F*_2,158_=12.14, *p*<.001, *η^2^_p_* =.13). Inter-groups comparisons and comparisons to normative values led to the characterization of Cluster 1 as normal cognition, Cluster 2 as probable executive MCI and Cluster 3 as probable amnestic and executive MCI.

**Conclusions:** Individuals with cognitive profiles consistent with probable executive MCI were identified using simple assessments of global cognition, memory and executive functions in a community-dwelling older population. As participants were not clinically diagnosed, further prospective studies are needed to determine the screening performance of this approach.

## 1 Introduction

Due to global population aging, dementia has become an increasingly important public health issue (Livingston et al., 2020). Among all forms of dementia, Alzheimer’s disease (AD) is the most prevalent, accounting for 60% to 70% of cases worldwide (World Health Organization, 2025). Consequently, considerable efforts have focused on the early identification of AD, with the aim of detecting signs and symptoms that may serve as predictive markers of disease progression (De Roeck et al., 2016). Mild Cognitive Impairment (MCI), a predementia stage, is characterized by a high risk of progression: 28.9% of MCI cases progress to AD and 21.9% to other types of dementia in population studies (Dubois and Albert, 2004; Mitchell and Shiri-Feshki, 2009). Early and accurate detection of MCI is therefore critical for implementing interventions that may delay or even prevent the onset of dementia.

MCI, the transitional stage between healthy aging and dementia, is defined by cognitive decline beyond expectations for age and education, while preserving autonomy in daily activities (Petersen et al., 2001). Its prevalence rises sharply with age, from 10.88% in adults aged 50–59 to 21.27% in those over 80 (Bai et al., 2022). Given the potential for distinct cognitive domains to be affected, MCI is heterogeneous (Anderson, 2019). It includes four subtypes based on memory involvement (amnestic *vs.* non-amnestic) and the number of affected domains (single *vs.* multiple) (Petersen, 2004). While the identification of MCI is of major importance, this heterogeneity complicates its detection and contributes to variability in cognitive assessment tools and clinical criteria (Portet, 2006; Albert et al., 2011; Díaz-Mardomingo et al., 2017). Additionally, identifying participants with early signs of probable MCI in community-dwelling older adults poses a significant challenge due to their high cognitive and functional reserves (Reuter-Lorenz and Park, 2014). High-functioning individuals often use compensatory mechanisms that can mask subtle cognitive decline on standard assessment tools until the pathology reaches a critical threshold (Corbo et al., 2023). Thus, sensitive, standardized, and multimodal assessments are essential to accurately classify MCI, especially in community-dwelling older adults (Klekociuk et al., 2014).

Current screening and diagnostic procedures for MCI rely on validated tools, with the Montreal Cognitive Assessment (MoCA) standing out as the preferred screening tool at the primary care level (Nasreddine et al., 2005; Abd Razak et al., 2019). With a cutoff score of 26, the MoCA achieves 90% sensitivity and 87% specificity in distinguishing MCI from cognitively normal older adults. However, the MoCA alone is insufficient for identifying the specific cognitive domains affected. The MoCA incorporates a delayed word list recall test that allows calculation of an independent score, the MoCA Memory Index Score (MIS). The MIS demonstrates good diagnostic performance, with 80% sensitivity and 69.1% specificity in differentiating individuals with amnestic MCI from cognitively normal older adults (Kaur et al., 2018). Notably, combining the total MoCA score with the MIS has been shown to predict conversion from MCI to AD with remarkable accuracy, as 90.5% of individuals with low baseline scores on both measures progressed to AD (Julayanont et al., 2014). These findings, observed in individuals already diagnosed with MCI, underscore the value of integrating multiple cognitive assessments to enhance diagnostic precision. Beyond memory, deficits in executive functioning are also prevalent in MCI populations (Corbo & Casagrande, 2022), reinforcing the need for a multidomain approach to improve subtype classification.

According to Diamond’s model, core Executive Functions (EFs) are cognitive flexibility, inhibition, and working memory (Diamond, 2013). In the context of MCI, cognitive flexibility and inhibition are particularly relevant (Corbo et al., 2024; Guarino et al., 2020). They are associated with greater difficulties in activities of daily living (Giovannetti et al., 2008; Aretouli and Brandt, 2010) and with both classification of cognitive impairment and conversion to AD in MCI patients (Guarino et al., 2019). Multivariate approaches have already demonstrated their effectiveness in this field. For instance, machine learning algorithms have accurately classified older adults according to their cognitive aging (cognitively healthy, MCI, or dementia) using only three cognitive domains: global cognitive status, memory, and executive functioning. In these models, performance on Part B of the Trail Making Test (TMT), which assesses cognitive flexibility, is a key variable (Weakley et al., 2015). Similarly, Chapman et al. (2011) demonstrated that adding executive functioning to memory-based models significantly improves the prediction accuracy of progression to AD in MCI patients. In their study, cognitive flexibility (assessed by the TMT) and inhibition (assessed by the Stroop Test) were the executive functions involved. While these studies focused on diagnosed MCI populations, we propose to apply a similar multivariate approach to a different context: community-dwelling older adults without a prior MCI diagnosis. From this perspective, combining a global cognitive screening tool such as the MoCA with simply complementary EF measures, namely cognitive flexibility (TMT) (Reitan, 1958) and inhibition (Victoria Stroop Test, V-ST) (Bayard et al., 2011), appears relevant. These neuropsychological tests are well-established, validated, easy to administer, and quick to complete. Moreover, the availability of normative standards for each test allows for the consideration of factors influencing cognitive aging (age, education level, gender) and the establishment of cutoff scores, further enhancing their utility in MCI screening for community-dwelling older adults.

In this paper, we propose a novel application of a multivariate classification method to identify distinct cognitive aging profiles in apparently cognitively healthy, autonomous older adults. This approach, inspired by methods previously validated in diagnosed MCI populations (*e.g.*, Weakley et al., 2015; Chapman et al., 2011), associates a clustering analysis based on scores from three well-established neuropsychological tests: MoCA, TMT, and V-ST. We hypothesize that (1) most participants will exhibit a normal cognitive aging profile; (2) a subgroup will present a profile consistent with probable MCI, characterized by lower MoCA scores; and (3) within this later subgroup, a significant proportion of the population will combine profiles consistent with probable MCI, poorer performance on executive function assessments, and lower MIS.

## 2 Methods

All data used in this study were extracted from the 13EVAL cluster randomized controlled trial (https://ClinicalTrials.gov identifier: NCT05625828). Pre-test and post-test assessment sessions included evaluations of functional capacities (*e.g*., spontaneous walking speed), cognitive functions, and cognitive-motor abilities. The data analyzed in the present work are derived exclusively from pre-test assessments.

### 2.1 Participants

The sample included 161 community-dwelling older adults (mean age of 76.05±6.36 years, range: 65–92). Among them, 85.71% were women (n=138). Participants had a mean Body Mass Index (BMI) of 24.82±3.66 kg.m-2 (range: 16.16–34.96). The average spontaneous walking speed was 1.11±0.18 m/s (range: 0.70–1.62). Participants were recruited by the French Rugby League Federation between September 13th, 2022, and April 24th, 2025. Inclusion criteria were: being 65 years of age or older, living in the community, being able to walk independently, and being able to follow instructions for testing. Exclusion criteria were: more than three falls in the last 12 months, musculoskeletal disorders impairing posture or gait, central or peripheral neurological disease (*e.g.,* previous stroke, Parkinson’s disease), psychiatric disorders, probable depression (Geriatric Depression Scale score > 10) (Yesavage and Sheikh, 1986), neurocognitive disorder (MoCA score<18) (Nasreddine et al., 2005), obesity (BMI>35), contraindication to practicing a physical activity, and involvement in another research protocol.

### 2.2 Ethics

The study was approved by a national ethic committee (CPP Île de France III – EUDRACT 2022-001517-38). All participants received written information about the nature and objectives of the study and provided written informed consent prior to participation. All procedures were conducted in accordance with the Declaration of Helsinki and complied with ethical standards, legal requirements and international norms.

### 2.3 Measures

#### 2.3.1 Global cognition

Global cognitive status was assessed using the MoCA (score range: 0–30) (Nasreddine et al., 2005). This instrument evaluates six cognitive domains: attention (6 points), executive functions (4 points), language (5 points), short-term memory (5 points), orientation (6 points), and visuospatial abilities (4 points). MoCA corrected score was adjusted for educational level, with one bonus point added for participants with ≤12 years of education, as recommended by Nasreddine et al., (2005).

#### 2.3.2 Memory

Short-term memory was assessed using the MoCA Memory Index Score (MIS), a sub-score derived from the delayed word list recall task incorporated in the MoCA. This task, based on Buschke et al.’s free and cued recall paradigm (1999), requires participants to recall five words after a 5-minute delay, during which they complete the remaining MoCA sections. The MIS is calculated independently of the global MoCA score by summing the correct responses from free delayed recall (weighted by 3), category-cued recall (weighted by 2), and multiple-choice cued recall (weighted by 1), yielding a score ranging from 0 to 15.

#### 2.3.3 Cognitive flexibility

Cognitive flexibility was assessed using the paper-and-pencil TMT (Reitan, 1958). This test consists of connecting the circled numbers and letters displayed on an A4 sheet as quickly as possible using a pencil. It includes two parts: Part A, in which participants connect numbers from 1 to 25 in ascending order, and Part B, in which participants alternate between numbers (1-12) and letters (A-L). Execution time was recorded for each part using a hand-held chronometer, and Delta-TMT was calculated as the difference between execution times in Part B and Part A, better reflecting cognitive flexibility than execution time in part B alone (Sánchez-Cubillo et al., 2009).

#### 2.3.4 Inhibition

Inhibition was assessed using the V-ST (Bayard et al., 2011), which comprises three conditions of increasing complexity: (i) Color condition (“Stroop-C”), consisting of colored dots; (ii) Word condition (“Stroop-W”), consisting of neutral words printed in colored ink (*e.g.,* “FOR”, *i.e.* “POUR” in French, printed in blue); and (iii) Interference condition (“Stroop-I”), consisting of color words printed in incongruent ink colors (*e.g.,* “GREEN”, *i.e.* “VERT” in French, printed in red). For each condition, participants were instructed to name the color of the ink of each stimulus as quickly as possible. Execution time was measured using a hand-held chronometer. The high interference ratio (Inter-Hi) was calculated according to the equation proposed by Bayard et al. (2011): Inter-Hi=Stroop-I/Stroop-C, better reflecting inhibition than execution time in Stroop-I alone (Van der Elst et al., 2006).

### 2.4 Data analysis

#### 2.4.1 Hierarchical clustering on principal component

To classify participants into distinct cognitive aging profiles, we used Hierarchical Clustering on Principal Components (HCPC), an unsupervised multivariate method (Husson et al., 2017). This approach combines Principal Component Analysis (PCA) to reduce dimensionality and Hierarchical Cluster Analysis (HCA) with Ward’s method to minimize intra-group inertia and maximize inter-group inertia (Ward, 1963). The clustering process followed four key steps: (1) PCA was performed to project individuals onto the component space, reducing data dimensionality; (2) HCA was applied to iteratively group individuals based on their similarity (*i.e.,* by bringing observations together one by one across the different components), generating a dendrogram; (3) The dendrogram was partitioned by identifying the largest increase in inertia (*i.e.*, the greatest aggregation index jump), which determined the optimal number of clusters, and (4) Cluster membership was refined using K-means consolidation to optimize within-cluster homogeneity. This procedure improves the stability and homogeneity of the final partition while preserving the number of clusters determined in the previous step.

Five active variables were included in the HCPC to capture the multi-dimensional nature of cognitive aging profiles: corrected MoCA score, MIS, Delta-TMT, Inter-Hi, and age. Age was included as an active variable because cognitive performance is inherently linked to chronological age, and accounting for it ensures that the identified profiles reflect age-related cognitive variations rather than being confounded by age differences. Analyses were conducted using the FactoMineR R package (v2.14) (Husson et al., 2026).

#### 2.4.2 Comparison with normative standards

To interpret individual performances on all conditions of the three neuropsychological tests, raw scores and indices were compared to age, education, and/or gender adjusted normative standards. First, norms validated from reference populations closely matching the sample included in the present study were prioritized (*i.e.,* French older adults). This was applied to the uncorrected MoCA score (*i.e.,* without the additional point for low educational level, following Larouche et al., (2016) guidelines), the Delta-TMT (Amieva et al., 2009), and the Inter-Hi (Bayard et al., 2011). The only exception was for the MIS, for which normative standard were only available from a Brazilian population (Apolinario et al., 2018). Second, to identify cognitive impairment, the “deficit” threshold recommended by Petersen, (2004) was used, defined as a performance falling below the 5^th^ percentile or a Z-score lower than -1.65. Third, standard clinical category labels were used across tests to facilitate interpretation while remaining consistent with the original validation studies: Very Superior, Superior, Average, Inferior, and Deficit. For example, the “Very Superior” category was not applied to the TMT because it was not defined in the study by Amieva et al. (2009).

Performances on the MoCA and the V-ST (in Z-scores) were classified as follows: >1.65 (very superior); [1.65 to 0.90[(superior); [0.90 to -0.90] (average);]-0.90 to -1.65] (inferior); and <-1.65 (deficit). For the TMT (in percentiles), the categories were: <25th (superior); [25th to 75th] (average);]75th to 95th] (inferior); and >95th (deficit).

### 2.5 Statistical analysis

Analyses were conducted using Jamovi® v2.6.17 (The jamovi project, 2025) and RStudio v2024.12.1+563 (R Core Team, 2025), with an alpha level of 0.05. Preliminary analyses were performed on dependent variables to check the normality of the distributions using the Kolmogorov-Smirnov test and the presence of outliers (Osborne, 2013). The Kolmogorov-Smirnov test was non-significant (p>.05) for all dependent variables.

One-way (Cluster) ANOVAs were performed to compare clusters with respect to their general characteristics, and (2) their performances in each variable used for HCPC (*i.e.,* MoCA, MIS, Delta-TMT, and Inter-Hi). For all ANOVAs, *post-hoc* comparisons were conducted using the Tukey correction when the main effect reached statistical significance. The effect sizes for all ANOVAs were reported using partial eta-squared (*η^2^_p_*) and using Cohen’s d for *post-hoc* analyses. The following thresholds were selected for reporting effect sizes: (i) for *η^2^_p_*.01-.06 small; .07-.14 medium; and >.14 large; and (ii) Cohen’s d 0.2-0.5 small; 0.6-0.8 medium; and >0.8 large (Cohen, 1988). Finally, Chi-square tests were performed to compare the global distribution of participants across normative categories among clusters, followed by pairwise comparisons.

## 3 Results

### 3.1 Cognitive assessment

More than half of the participants (53.42%, n=86) had completed at least 12 years of education. Descriptive results for the cognitive tests are presented in Table 1. Notably, more than half of the participants (54.04%, n=87) obtained a score below the recommended cutoff of 26/30 on the MoCA, suggesting probable MCI.

**Table 1.** Results of the cognitive assessment.

| Test | | | Mean ± SD | Range |
| --- | --- | --- | --- | --- |
| Name | Function | Variable | (N = 161) | [min; max] |
| MoCA | Global cognition | MoCA (score/30) | 24.94 ± 2.95 | [18; 30] |
| | Memory | MIS (score/15) | 10.71 ± 3.99 | [0; 15] |
| TMT | Cognitive flexibility | TMT-A (s) | 39.59 ± 14.84 | [20.80; 96.00] |
| | | TMT-B (s) | 93.72 ± 44.11 | [35.70; 251.00] |
| Victoria Stroop Test | Inhibition | Stroop-C (s) | 15.05 ± 4.04 | [8.00; 32.50] |
| | | Stroop-W (s) | 19.76 ± 4.85 | [12.60; 37.50] |
| | | Stroop-I (s) | 35.22 ± 11.06 | [16.70; 94.00] |

SD = Standard Deviation; MoCA = Montreal Cognitive Assessment; MIS = Memory Index Score; TMT-A = Trail Making Test, part A; TMT-B = Trail Making Test, part B; Stroop-C = Color condition; Stroop-W= Word condition; Stroop-I = Interference condition.

### 3.2 Cluster Classification Based on Principal Components

The hierarchical clustering performed on the principal components revealed a three-cluster structure. The dendrogram (Ward’s method) showed a marked increase in fusion distance beyond the three-cluster solution, supporting its selection (see Supplementary Figure 1). Cluster 1 appeared highly distinct, merging with the other clusters only at a high level of dissimilarity, whereas Clusters 2 and 3 showed greater proximity, suggesting partially shared characteristics. The distribution of individuals across the three PCA dimensions according to cluster membership is presented in Figure 1.

**Figure 1.**
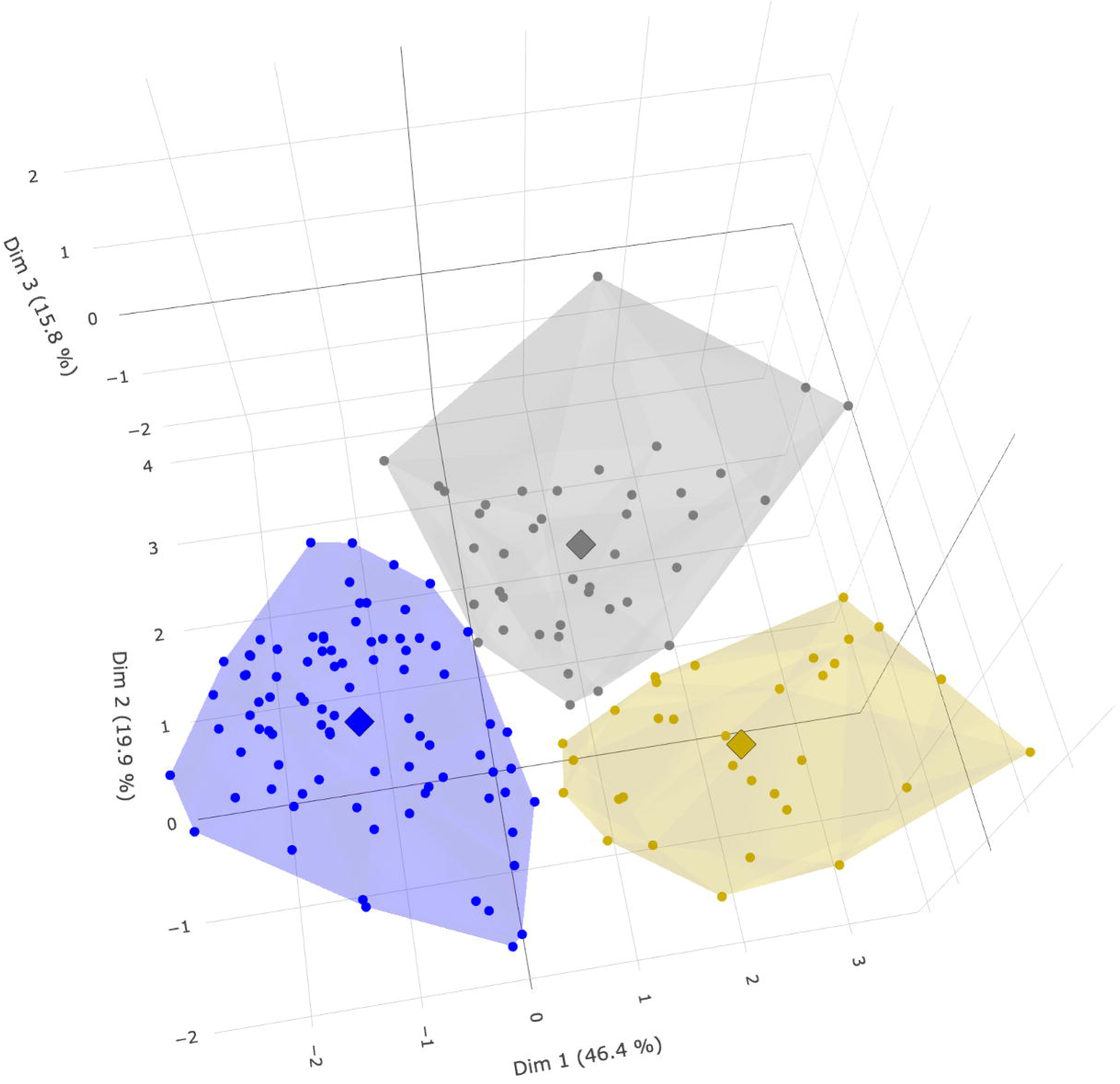
Individuals grouped by clusters in the three dimensions determined by the principal component analysis and percentage of variance explained by each dimension. Dim: dimension, Blue: Cluster 1, Grey: Cluster 2, Yellow: Cluster 3, Points: individual participants, Diamond shapes: mean value of each cluster.

### 3.3 Characteristics and Differences among Identified Clusters

Regarding age, a significant cluster effect was found (*F*_2,158_=78.56, *p*<.001, *η_2p_*=.50, large effect): *post-hoc* comparisons showed a significant difference between clusters 1 and 2 (*t*_158_=-10.84, *p*<.001, *d*=-1.88, large effect) and between clusters 1 and 3 (*t*_158_=-9.24, *p*<.001, *d*=-2.18, large effect). Regarding MoCA score, a significant cluster effect was found (*F*_2,158_=96.07, *p*<.001, *η^2^_p_* =.55, large effect): *post-hoc* comparisons showed a significant difference between clusters 1 and 2 (*t*_158_=8.85, *p*<.001, *d*=1.53, large effect) and clusters 1 and 3 (*t*_158_=12.82, *p*<.001, *d*=3.03, large effect) and between clusters 2 and 3 (*t*_158_=6.02, *p*<.001, *d*=1.49, large effect). Regarding MIS, a significant cluster effect was found (*F*_2,158_=81.48, *p*<.001, *η^2^_p_* =.51, large effect): *post-hoc* comparisons showed a significant difference between clusters 1 and 2 (*t*_158_=4.25, *p*<.001, *d*=0.74, medium effect) and between clusters 1 and 3 (*t*_158_=12.76, *p*<.001, *d*=3.01, large effect) as well as between clusters 2 and 3 (*t*_158_=9.18, *p*<.001, *d*=2.27, large effect). Regarding Delta-TMT, a significant cluster effect was found (*F*_2,158_=52.25, *p*<.001, *η_2p_* =.40, large effect): *post-hoc* comparisons showed a significant difference between clusters 1 and 2 (*t*_158_=-5.42, *p*<.001, *d*=-0.94, large effect) and between clusters 1 and 3 (*t*_158_=-9.89, *p*<.001, *d*=-2.23, large effect) as well as between clusters 2 and 3 (*t*_158_=-5.63, *p*<.001, *d*=-1.39, large effect). Regarding Inter-Hi, a significant cluster effect was found (*F*_2,158_=12.14, *p*<.001, *η_2p_* =.13, medium effect): *post-hoc* comparisons showed significant differences between clusters 1 and 2 (*t*_158_=-4.64, *p*<.001, *d*=-0.80, large effect) and between clusters 1 and 3 (*t*_158_=-2.97, *p*<.05, *d*=-0.70, medium effect). The characteristics of the five variables used in the HCPC for each cluster are presented in Table 2. The statistical differences between clusters are indicated in Figure 2.

**Table 2.** Characteristics of the five variables used in the HCPC for each cluster.

| Variable | Cluster |  |  |
| --- | --- | --- | --- |
|  | Cluster 1 (N=82) | Cluster 2 (N=56) | Cluster 3 (N=23) |
| Age | 71.86±4.39<br>[65.00; 81.08] | 80.42±4.61<br>[72.08; 92.50] | 81.17±4.97<br>[68.50; 92.33] |
| MoCA | 26.87±1.97<br>[21; 30] | 23.80±2.07<br>[18; 29] | 20.83±1.92<br>[18; 24] |
| MIS | 12.65±2.74<br>[3; 15] | 10.57±2.81<br>[3; 15] | 4.17±3.08<br>[0; 10] |
| Delta-TMT | 35.68±19.95<br>[5.70; 89.44] | 61.94±30.39<br>[5.00; 136.10] | 100.94±42.93<br>[37.30; 192.50] |
| Inter-Hi | 2.16±0.49<br>[1.20; 3.49] | 2.68±0.76<br>[1.48; 5.28] | 2.61±0.82<br>[1.42; 4.95] |

Values are mean ± standard deviation [Minimum; Maximum]
MOCA = Montreal Cognitive Assessment.
MIS= Memory index score.
Delta-TMT =Trail making test (Part B - Part A).
Inter-Hi=The high interference ratio (Stroop-I/Stroop-C).

**Figure 2.**
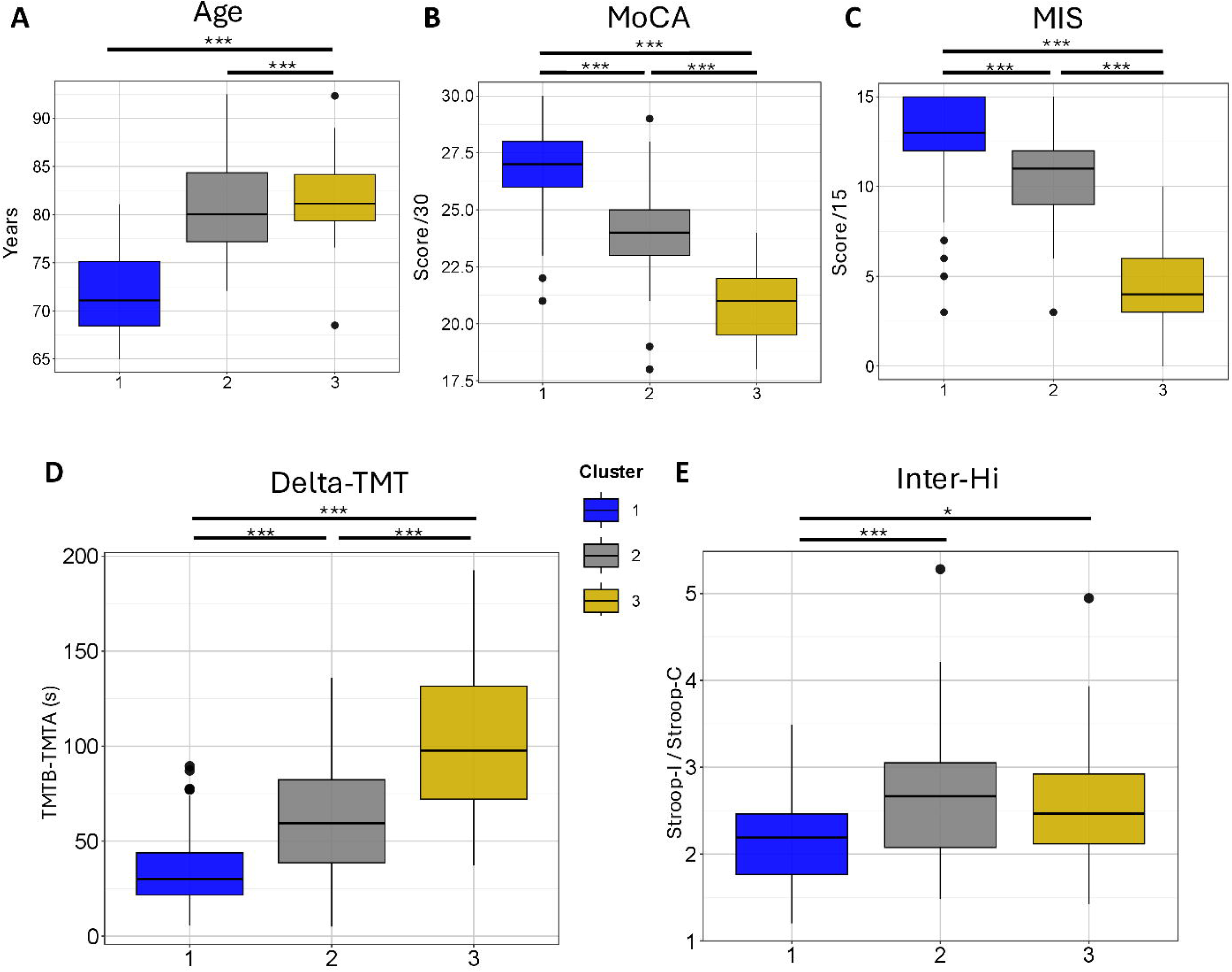
Boxplots representing post-hoc comparisons between clusters from hierarchical clustering on principal component analysis.

### 3.4 Distribution of Normative Cognitive Classifications Across Clusters

The distribution of normative categories revealed distinct performance patterns for the MoCA, Delta-TMT, and Inter-Hi across the three clusters identified by hierarchical clustering. The distribution of normative categories for each cluster and each neuropsychological test condition is presented in supplementary data (Supplementary Table 1). Attention was particularly given to the variables included in the HCPC to characterize each cluster.

In **Cluster 1**, MoCA scores were mainly classified as average (56.10%, n=46), followed by superior (18.29%, n=15), very superior (10.98%, n=9) and inferior (10.98%, n=9). MIS scores were mainly classified as superior (54.88%, n=45) followed by average (39.02%, n=32), inferior (3.66%, n=3) and very superior (2.44%, n=2) scores. Delta-TMT performance was mainly classified as superior (65.85%, n=54), followed by average (23.17%, n=19) and inferior (10.98%, n=9). Inter-Hi performance was mainly classified as average (54.88%, n=45), followed by superior (29.27%, n=24), very superior (9.76%, n=8), and inferior (6.10%, n=5).

In **Cluster 2**, MoCA scores were mainly classified as average (50.00%, n=28), followed by inferior (23.21%, n=13), deficit (17.86%, n=10), superior (7.19%, n=4) and very superior (1.79%, n=1). MIS scores were mainly classified as average (55.36%, n=31) followed by superior (37.50%, n=21), very superior (5.36%, n=3) and inferior (1.79%, n=1). Delta-TMT performance was mainly classified as average (50.00%, n=28), followed by superior (32.14%, n=18), inferior (14.29%, n=8) and deficit (3.57%, n=2). Inter-Hi performance was mainly classified as average (57.14%, n=32) followed by superior (21.43%, n=12), inferior (14.29%, n=6), deficit (5.36%, n=3), and very superior (1.79%, n=1).

In **Cluster 3**, MoCA scores were mainly classified as deficit (39.13%, n=9), followed by inferior (34.78%, n=8) and average (26.09%, n=6). MIS scores were mainly classified as average (60.87%, n=14) followed by inferior (21.74%, n=5) and deficit (17.39%, n=4). Delta-TMT performance was equally classified as average and inferior (34.78%, n=8), followed by deficit (17.39%, n=4), and superior (13.04%, n=3). Inter-Hi scores were mainly classified as average (65.22%, n=15), followed by very superior (13.04%, n=3), equally by superior and inferior (8.70%, n=2), and then deficit (4.35%, n=1).

The overall distribution of normative categories between the three Clusters is presented in Figure 3. For MoCA, this overall distribution of normative categories differed significantly across the Clusters (*χ*^2^_8_=40.20, p<.001). Pairwise comparisons showed a significantly different distribution of normative classes between Clusters 1 and 2 (*χ*^2^_4_ =17.36, *p*<.005) and between Clusters 1 and 3 (*χ*^2^_4_ =36.06, *p*<.001). A similar overall result was found for the MIS (*χ*^2^_8_ =54.73, *p*<.001). Pairwise comparisons showed a significantly different distribution of normative classes between Clusters 1 and 3 (*χ*^2^_4_ =37.11, *p*<.001) and between Clusters 2 and 3 (*χ*^2^_4_ =28.23, *p*<.001). Similar overall result was found for Delta-TMT (*χ*^2^_6_ =42.84, *p*<.001). Pairwise comparisons showed a significantly different distribution of normative classes between Clusters 1 and 2 (*χ*^2^_3_ =17.51, *p*<.001), between Clusters 1 and 3 (*χ*^2^_3_ =30.72, *p*<.001) and between Clusters 2 and 3 (*χ*^2^_3_ =10.55, *p*<.05). For Inter-Hi, no significant overall difference was found (*χ*^2^_8_=14.31, *p*=.807) although the overall test was not significant, an exploratory pairwise comparison was conducted and suggest a significantly different distribution of normative classes between Clusters 1 and 2 (*χ*^2^_4_ =10.82, *p<*.05). However, this result should be interpreted with caution due to the non-significance of the overall Inter-Hi model.

**Figure 3.**
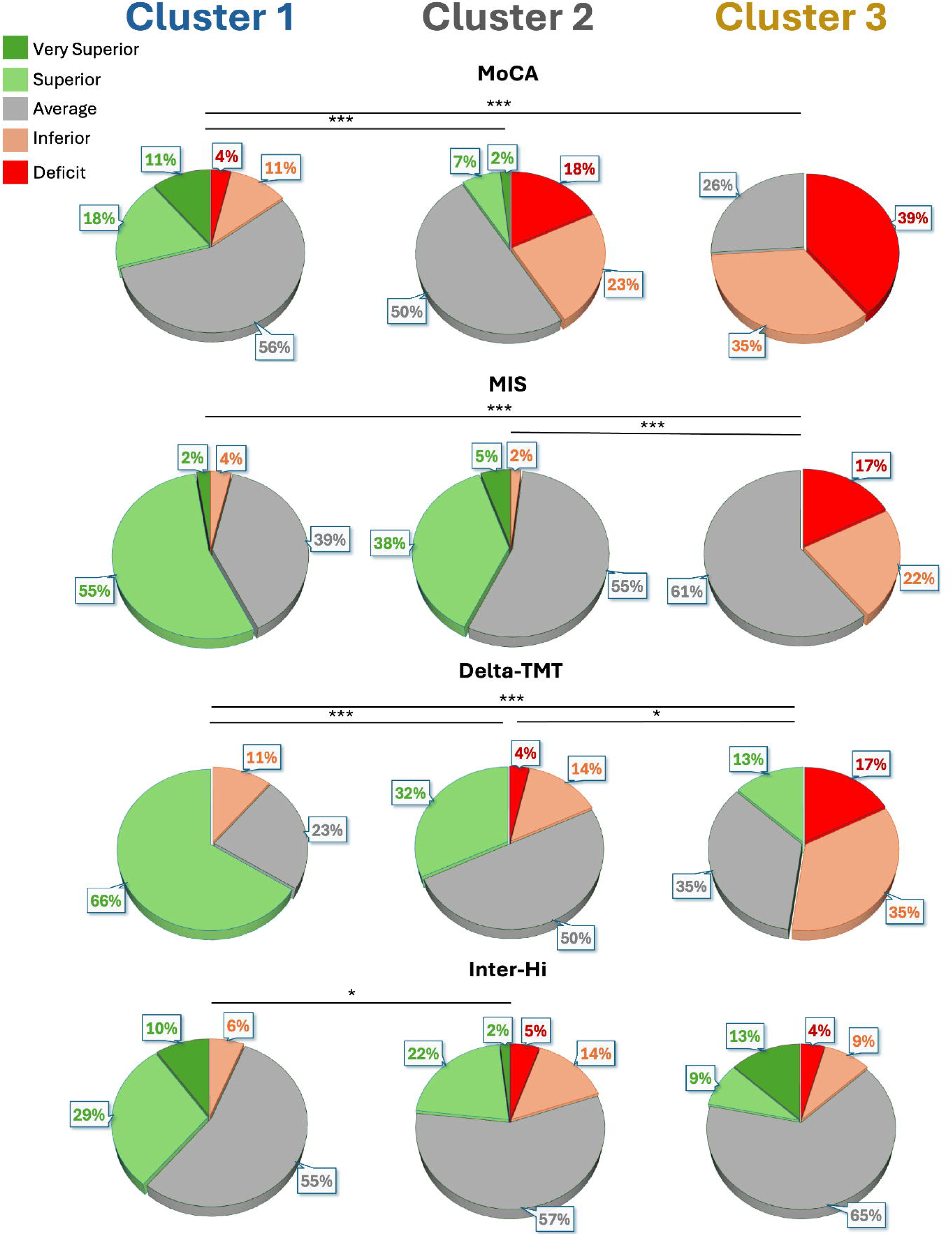
Proportions of each normative categories by cluster and pairwise comparisons based on Chi-squared test.

## 4 Discussion

The aim of this study was to identify different cognitive aging profiles in a population of apparently cognitively healthy, community-dwelling older adults using a multivariate, unsupervised classification approach (HCPC) based on the MoCA total score, the MoCA Memory Index Score, and simple executive function assessments (TMT and V-ST). This approach, previously validated in diagnosed MCI populations, revealed three distinct cognitive profiles: a profile consistent with normal cognitive aging, a profile characterized by predominant executive difficulties, and a profile combining memory and executive impairments consistent with probable multidomain MCI. Importantly, these findings suggest that combining global cognition, memory, and EF assessments provides a more informative and sensitive characterization of cognitive vulnerability than relying on a global screening score alone, even in the absence of clinical diagnoses.

Interestingly, prior to clustering, 54.04% of the overall sample presented a MoCA score below the standard threshold of <26, suggesting an increased likelihood of cognitive impairment. This proportion appears substantially higher than typical epidemiological estimates of MCI (Bai et al., 2022). One possible explanation may relate to the high sensitivity and the relative lower specificity of the MoCA reported in the recent Cochrane review (Davis et al., 2021). Hence, applying a strict cutoff of <26 may generate false positives, especially in a sample where half of the participants had more than 12 years of education (Kaur et al., 2018). Alternatively, participants appeared relatively preserved functional capacities, as reflected by an average gait speed (1.11 m/s) well above frailty thresholds reported in geriatric literature (Montero-Odasso et al., 2005; Houles et al., 2010). This functional robustness may explain why a substantial proportion of the sample scored below the MoCA threshold without showing any marked limitations in daily functioning. This interpretation is consistent with the “scaffolding” theory, according to which high-functioning individuals may compensate for early neural decline until compensatory mechanisms become insufficient (Reuter-Lorenz and Park, 2014). Together, these findings support the value of a multi-component clustering approach to more accurately characterize cognitive profiles rather than relying solely on a global cut-off score. In addition, to overcome limitations associated with raw scores, current clinical recommendations emphasize the use of age- and education-adjusted normative data, particularly when distinguishing between MCI subtypes (Petersen, 2004; Petersen et al., 2014).

Clustering analysis revealed three distinct cognitive profiles in this tested population of apparently cognitively healthy community-dwelling older adults. Participants in Cluster 1 (n=82) demonstrated a preserved cognitive profile, characterized by superior or average performances across the MoCA, MIS, TMT, and V-ST. This group aligns with the concept of “successful aging” (Rowe and Kahn, 1987). Their preserved cognitive abilities may reflect relatively high cognitive and functional reserves (Reuter-Lorenz and Park, 2014). Given that participants were recruited within the context of a physical exercise intervention trial, it is plausible that their baseline physical engagement and autonomy contributed to this preserved cognitive profile. In contrast, Clusters 2 (n=56) and 3 (n=23) exhibited cognitive profiles consistent with different probable MCI subtypes. Cluster 2 showed significantly lower MIS than Cluster 1 (medium effect) but higher than Cluster 3 (large effect), and significantly lower V-ST performance than Cluster 1 (large effect). Cluster 3 is particularly worth highlighting as it displayed a profile consistent with probable amnestic multiple-domain MCI (amdMCI), characterized by combined alterations in memory and EF. Individuals presenting amnesic impairment combined with executive dysfunction are considered at higher risk of progression to dementia, particularly AD (Chapman et al., 2011; Zhuang et al., 2021). In the early stages of MCI, neuroimaging and behavioral models suggest that preserved frontal and executive networks may support compensatory mechanisms in response to hippocampal dysfunctions, which primarily manifest as memory impairment (Dannhauser et al., 2008; Clément et al., 2010). As disease progresses, these frontal executive networks are also affected and may become less able to sustain this compensation marking a critical transition toward dementia (Kim et al., 2016). This framework may provide one possible explanation for the concomitant alterations in memory and EF observed in Cluster 3.

However, age-related deficit in EF may differentially affect specific executive sub-domains. Results of the present study highlight differences in the discriminative ability of the EF assessments used to characterize cognitive profiles. While both cognitive flexibility, reflected by Delta-TMT, and inhibition, reflected by Inter-Hi, differed significantly across clusters, the magnitude of these effects varied considerably. Cognitive flexibility showed a large cluster effect (*η^2^_p_* =.40) and *post-hoc* analyses revealed that it differentiated the profile consistent with probable amdMCI profile (Cluster 3) from both the normal cognition profile (Cluster 1) and the profile consistent with probable amnestic single-domain MCI (Cluster 2). In contrast, inhibition yielded a smaller medium-sized group effect (*η^2^_p_* =.13). Although inhibition differentiated the normal cognition profile from the two probable MCI-related profiles, its overall discriminative capacity appeared lower. These findings suggest that, in this population of community-dwelling older adults, a decline in cognitive flexibility may represent a more sensitive marker of multiple-domain MCI than deficits in inhibition (Corbo et al., 2024). One possible explanation is that task-switching capacity (set-shifting) may be more strongly affected than inhibitory processes in individuals with high functional reserve but presenting early cognitive changes. These findings extend previous work emphasizing the importance of selecting change-sensitive EF assessments for the early identification of cognitive decline (De Roeck et al., 2019). Consequently, combining flexibility measures with global cognitive screening may improve characterization of heterogeneous cognitive aging profiles and contribute to the identification of executive dysfunction in individuals presenting higher risk of progression to AD.

## 5 Limitations & Perspectives

Several limitations should be considered when interpreting these findings. First, there is a significant age difference between the clusters, with participants in Cluster 3 being significantly older than the others. It is therefore difficult to fully disentangle cognitive decline from age-related changes. Second, participants were community-dwelling older adults without a formal clinical diagnosis of MCI, meaning the identified clusters should be interpreted as cognitive profiles consistent with probable MCI rather than clinically confirmed MCI subtypes. Third, most participants were women (85.71%), which may limit generalization of the results to older men. Finally, although paper-and-pencil neuropsychological assessments remain valuable screening tools, high-functioning older adults may still perform within normative ranges due to high functional reserve. Future studies could therefore incorporate more ecological approaches including cognitive-motor interference and locomotor performance. Complex walking tests, such as dual-task paradigms, may provide complementary and potentially more sensitive indicators of early cognitive vulnerability in highly functional older adults (Grosboillot et al., 2024; Mauti et al., 2026).

## 6 Conclusions

The present findings suggest that a multivariate approach combining simple, rapid, and widely accessible neuropsychological assessments aligns with the growing need for sensitive and scalable screening tools but also underscores the potential of multivariate methods to capture the heterogeneity of cognitive aging in real-world settings. While most participants exhibited preserved cognitive or isolated cognitive alterations, the clustering approach identified a subgroup displaying a profile consistent with probable amdMCI. Notably, the findings suggest that cognitive flexibility, assessed using the TMT, showed greater discriminative capacity than cognitive inhibition assessed using the V-ST for distinguishing this profile. Identifying this probable amdMCI profile may be clinically relevant, as it has been associated with a higher risk of progression toward dementia. Individuals presenting such profiles may benefit from further clinical assessment using more comprehensive diagnostic procedures. Given that participants in this study were autonomous and not previously diagnosed, these findings highlight the potential presence of underrecognized cognitive vulnerabilities in community-dwelling older adults. Future research should extend these findings by incorporating more ecological and dynamic approaches, including complex walking and cognitive-motor dual-task paradigms, to improve the characterization of early cognitive changes in highly functional individuals.

## Supporting information

Distribution of normative classes for the three cluster in each condition of all neuropsychological tests

## 8 Abbreviations

AD: Alzheimer’s disease
amdMCI: amnestic multiple-domain MCI
BMI: Body Mass Index
EF: Executive Functions
HCA: Hierarchical Cluster Analysis
HCPC: Hierarchical Clustering on Principal Components
Inter-Hi: high interference ratio
MCI: Mild Cognitive Impairment
MIS: Memory Index Score
MoCA: Montreal Cognitive Assessment
PCA: Principal Component Analysis
TMT: Trail Making Test
V-ST: Victoria Stroop Test.

## 9 Acknowledgments

The authors would like to express their sincere gratitude to the Ligue Auvergne Rhône Alpes de Rugby à XIII for its financial and material support, as well as for its invaluable assistance in recruiting participants for this study. We would also like to thank its president, Mr. Jacques CAVEZZAN, for his enthusiasm and unconditional support throughout Rafael MAUTI’s doctoral project. Furthermore, the authors would like to extend their warmest thanks to the Grants Office of the Vaud Canton University of Teacher Education for its support throughout the publication process. Finally, we would also like to thank geriatrician and neurologist Antoine Garnier-Crussard (MD-PhD) of the Clinical and Research Memory Center at the Hospices Civils de Lyon for his careful review of this manuscript.

## 10 Author contributions

RM: Data curation, Formal analysis, Investigation, Methodology, Visualization, Writing – original draft, Writing – review & editing

CC: Writing – original draft, Writing – review & editing

AG: Writing – original draft, Writing – review & editing

AP: Investigation, Writing – review & editing

PF: Funding acquisition, Supervision, Writing – original draft, Writing – review & editing

PC: Conceptualization, Data curation, Funding acquisition, Project administration, Supervision, Writing – original draft, Writing – review & editing

## 12 Data availability statement

The data analyzed in this study is subject to the following licenses/restrictions: the data used in this study is available for research purposes on reasonable request to P.F. Requests to access these datasets should be directed to Patrick Fargier

---

**Supplementary Figure 1** Dendrogram of Hierarchical Clustering based on the Ward’s criterion. The height of the branches indicates the dissimilarity between clusters. The dendrogram was partitioned (black horizontal line) to maximize the distance between nodes. Cluster solution is indicated by three colored groups.

