## Supplementary material for "A clustering approach based on neuropsychological testing to detect early signs of probable MCI among community-dwelling older adults": Distribution of normative classes for the three cluster in each condition of all neuropsychological tests

**S1 Table 1: Distribution of normative classes for the three cluster in each condition of all neuropsychological tests.**

| Test | Condition | Category | Cluster 1 (n=82) | | Cluster 2 (n=56) | | Cluster 3 (n=23) | | $\chi^2_8$ ($p$) |
|---|---|---|---|---|---|---|---|---|---|
| | | | n | % | n | % | n | % | |
| MoCA | MoCA Score | Deficit | 3 | 3.66 | 10 | 17.86 | 9 | 39.13 | 40.20 |
| | | Inferior | 9 | 10.98 | 13 | 23.21 | 8 | 34.78 | (<.001) |
| | | Average | 46 | 56.10 | 28 | 50.00 | 6 | 26.09 | |
| | | Superior | 15 | 18.29 | 4 | 7.14 | 0 | 0.00 | |
| | | Very Superior | 9 | 10.98 | 1 | 1.79 | 0 | 0.00 | |
| | MIS | Deficit | 0 | 0.00 | 0 | 0.00 | 4 | 17.39 | 54.73 |
| | | Inferior | 3 | 3.66 | 1 | 1.79 | 5 | 21.74 | (<.001) |
| | | Average | 32 | 39.02 | 31 | 55.36 | 14 | 60.87 | |
| | | Superior | 45 | 54.88 | 21 | 37.50 | 0 | 0.00 | |
| | | Very Superior | 2 | 2.44 | 3 | 5.36 | 0 | 0.00 | |
| TMT | TMT-A | Deficit | 0 | 0.00 | 0 | 0.00 | 0 | 0.00 | *17.40 |
| | | Inferior | 3 | 3.66 | 5 | 8.93 | 1 | 4.35 | (<.005) |
| | | Average | 10 | 12.20 | 9 | 16.07 | 11 | 47.83 | |
| | | Superior | 69 | 84.15 | 42 | 75.00 | 11 | 47.83 | |
| | TMT-B | Deficit | 0 | 0.00 | 0 | 0.00 | 2 | 8.70 | $43.88 |
| | | Inferior | 3 | 3.66 | 4 | 7.14 | 5 | 21.74 | (<.001) |
| | | Average | 20 | 24.39 | 30 | 53.57 | 13 | 56.52 | |
| | | Superior | 59 | 71.95 | 22 | 39.29 | 3 | 13.04 | |
| | Delta-TMT | Deficit | 0 | 0.00 | 2 | 3.57 | 4 | 17.39 | $42.84 |
| | | Inferior | 9 | 10.98 | 8 | 14.29 | 8 | 34.78 | (<.001) |
| | | Average | 19 | 23.17 | 28 | 50.00 | 8 | 34.78 | |
| | | Superior | 54 | 65.85 | 18 | 32.14 | 3 | 13.04 | |
| V-ST | Stroop-C | Deficit | 4 | 4.88 | 2 | 3.57 | 1 | 4.35 | 13.32 |
| | | Inferior | 7 | 8.54 | 3 | 5.36 | 3 | 13.04 | (.101) |
| | | Average | 48 | 58.54 | 24 | 42.86 | 15 | 65.22 | |
| | | Superior | 21 | 25.61 | 19 | 33.93 | 3 | 13.04 | |
| | | Very Superior | 2 | 2.44 | 8 | 14.29 | 1 | 4.35 | |
| | Stroop-W | Deficit | 5 | 6.10 | 2 | 3.57 | 2 | 8.70 | 8.15 |
| | | Inferior | 9 | 10.98 | 6 | 10.71 | 5 | 21.74 | (.419) |
| | | Average | 56 | 68.29 | 31 | 55.36 | 12 | 52.17 | |
| | | Superior | 9 | 10.98 | 13 | 23.21 | 3 | 13.04 | |
| | | Very Superior | 3 | 3.66 | 4 | 7.14 | 1 | 4.35 | |
| | Stroop-I | Deficit | 0 | 0.00 | 1 | 1.79 | 1 | 4.35 | 13.31 |
| | | Inferior | 1 | 1.22 | 6 | 10.71 | 2 | 8.70 | (.093) |
| | | Average | 53 | 64.63 | 26 | 46.43 | 13 | 56.52 | |
| | | Superior | 17 | 20.73 | 18 | 32.14 | 4 | 17.39 | |
| | | Very Superior | 11 | 13.41 | 5 | 8.93 | 3 | 13.04 | |
| | Inter-Low | Deficit | 1 | 1.22 | 3 | 5.36 | 1 | 4.35 | 10.92 |
| | | Inferior | 3 | 3.66 | 8 | 14.29 | 2 | 8.70 | (.206) |
| | | Average | 56 | 68.29 | 33 | 58.93 | 12 | 52.17 | |
| | | Superior | 16 | 19.51 | 7 | 12.50 | 7 | 30.43 | |
| | | Very Superior | 6 | 7.32 | 5 | 8.93 | 1 | 4.35 | |
| | Inter-Hi | Deficit | 0 | 0.00 | 3 | 5.36 | 1 | 4.35 | 14.31 |
| | | Inferior | 5 | 6.10 | 8 | 14.29 | 2 | 8.70 | (.807) |
| | | Average | 45 | 54.88 | 32 | 57.14 | 15 | 65.22 | |
| | | Superior | 24 | 29.27 | 12 | 21.43 | 2 | 8.70 | |
| | | Very Superior | 8 | 9.76 | 1 | 1.79 | 3 | 13.04 | |

*Note*. "Deficit" class was defined for MoCA, MIS and V-ST by a Z-score inferior or equal to -1.65 and for the TMT by a performance below or equal to the 5th percentile. *Degrees of freedom for TMT-A are 4; $Degrees of freedom for TMT-B and Delta TMT are 6.
