## Supplementary figures and images for "A clustering approach based on neuropsychological testing to detect early signs of probable MCI among community-dwelling older adults"

### Supplemental Data 1

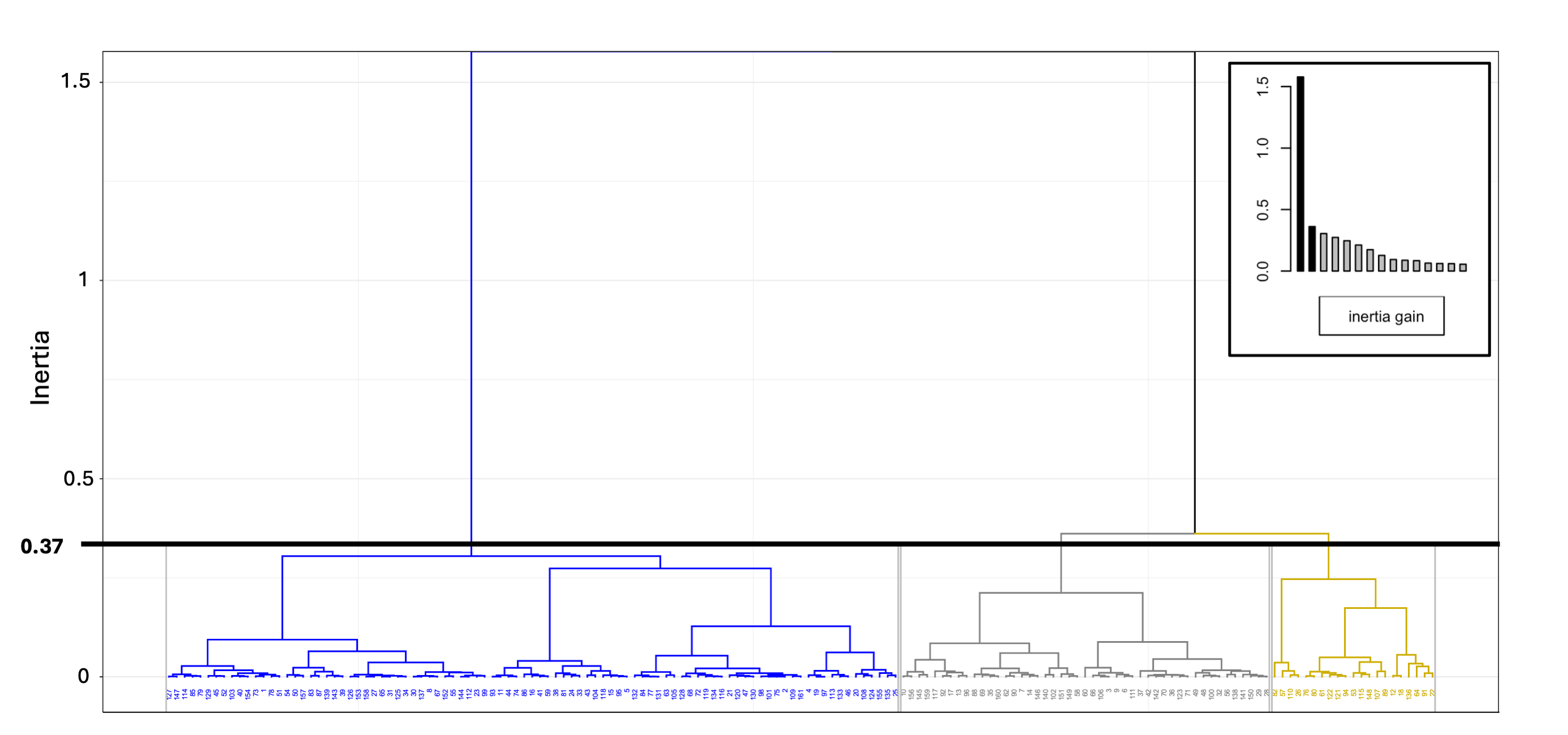
